# Safety and Efficacy of Navigational Bronchoscopy with Archimedes Virtual Bronchoscopic Platform in Patients with Advanced Pulmonary Disease

**DOI:** 10.1101/2022.01.22.22269681

**Authors:** Steven R. Verga, Robert Marron, Gerard J. Criner, Clinton Veselis, Chandra Dass, Hauqing Zhao

## Abstract

**Background:** Patients with advanced lung disease who present with suspicious pulmonary nodules (SPNs) undergoing transthoracic needle aspiration pose a diagnostic challenge given risk for complications. Virtual bronchoscopic navigation (VBN) is an alternative means to biopsy patients.

**Study Design:** The study was a retrospective chart review of all patients who underwent VBN with the Archimedes platform. Demographic information, radiographic evidence of parenchymal lung disease and pulmonary function testing were gathered. Peri-procedure complications and clinical data on biopsy results were gathered to assist in determining both adequate tissue sample (defined if adequate specimen was obtained as per pathology/cytology report) and diagnostic yield (defined as if after further testing/CT imaging from 1 year post-procedure the diagnosis did not change). Univariate analysis was performed to determine the impact of clinical factors on diagnostic yield.

**Results:** Ninety-six patients with 110 nodules underwent VBN. Eight-eight patients (92%) had radiographic emphysema with 56% having moderate to severe. Fourteen patients (17%) had radiographic ILD. Adequate tissue sample at time of VBN was 93%. Overall diagnostic yield in 80 patients (excluding 16 patients who were initially benign/non-diagnostic that were lost to follow-up) was 72%. Overall complication rate was 7% with 1-pneumothorax (1%), 3-significant bleeding and 3-hospitalized for respiratory failure.

**Conclusion:** In our cohort of patients the diagnostic yield and complication rate were comparable to other studies where there were fewer patients with advanced lung disease. This data suggests that VBN-guided bronchoscopic biopsy of SPNs is a viable diagnostic option with an acceptable safety profile.

## Introduction

Lung cancer is the leading cause of cancer-related deaths (18%) worldwide with 70% of patients presenting with an advanced stage at time of diagnosis.^1,2^ The 5-year survival of stage I non-small cell lung cancer (NSCLC) is more promising at 70-90% versus stage IV disease where 1-year survival is 15-19%.^3^ Accurate and timely diagnosis of suspicious pulmonary nodules (SPNs) that have malignant potential is crucial to avoid delays in diagnosis that lead to further disease progression and potentially lack of curative treatment options.^4,5^

Two large, randomized control trials utilizing low-dose computed tomography (LDCT) for lung cancer screening (e.g., National Lung Screening Trial [NLST] and NELSON trial) have reported improved survivals. With more widespread adoption of LDCT for lung cancer screening more incidental pulmonary nodules will be discovered.

Common diagnostic methods for biopsying pulmonary nodules are video-assisted thoracic surgery (VATS) (if highly suspicious for malignancy), CT-guided transthoracic needle aspiration (TTNA), or advanced bronchoscopy. Advanced bronchoscopic options include both radial and linear endobronchial ultrasound (EBUS), electromagnetic navigational bronchoscopy (ENB), virtual bronchoscopic navigation (VBN), CT-guided bronchoscopy, ultrathin bronchoscopy, robotic guide sheaths, and trans-parenchymal tunneling. Of these methods TTNA has a sensitivity between 74-90% with more studies reporting sensitivities closer to 90%, however potential complications include: pneumothorax (PTX) (as high as 35% with increased risk with emphysema), tension PTX, tumor seeding at site of biopsy route, hemothorax, hemoptysis, hemorrhage and air embolism.^6-8^ With TTNA and surgical resection having significant morbidity, bronchoscopic guided biopsy of SPNs may provide an alternative means with an overall lower risk of pneumothorax (1.5-3.1%).^9^ This is especially important in patients with SPNs and moderate to severe underlying lung disease where the risk of pneumothorax and complications of respiratory failure are greater.

Multiple retrospective studies reporting on use of VBN for the biopsy of pulmonary nodules report a diagnostic yield range of 71-88%, however, most of these studies were single center with limited follow up for indeterminate or non-diagnostic specimens.^10,11^ More recently the AQuIRE and NAVIGATE studies, both multi-center prospective studies utilizing VBN showed a wide diagnostic yield range of 39% (VBN alone without radial-EBUS) to 73% which accounted for delay in diagnosis due to need for subsequent work-up.^12,13^

The majority of our patients who are referred for diagnosis of SPNs have some form of advanced lung disease thus limiting the ability of patients to undergo CT-guided TTNA for SPN due to increased risk of complications. We performed an observation retrospective single center analysis of all patients undergoing VBN to determine overall diagnostic yield and safety in those with advanced underlying lung disease.

## Methods

### Study Design and Patient Enrollment

A retrospective chart review was performed on all consecutive patients who underwent VBN via Archimedes System (Broncus Medical, Inc., Redwood CA) platform to biopsy SPNs at Temple University Hospital (TUH) from December 2016 to October 2019 utilizing Olympus BF-P190 diagnostic bronchoscope. TUH is a large referral center due to our lung transplant program resulting in large percentage of patients referred for VBN having advanced lung disease. Data were obtained directly from the Archimedes VBN platform as well as the electronic medical record. Chart reviews were performed on every patient from the date of the procedure to one year afterward in order to determine the outcome for nodules with indeterminate or benign diagnoses; this was the same follow-up period as the NAVIGATE trial.^13^ The study was approved by the Institutional Review Board (Protocol ID: 26644).

### Archimedes VBN Platform

The Archimedes system is a VBN platform that integrates CT imaging with fused fluoroscopy to provide three-dimensional model to mark SPN(s) which the software then calculates several pathway(s) to the target SPN(s).^14^ This visually guides the user’s diagnostic bronchoscope (2mm working channel) along the selected pathway to the SPN of interest to utilize the biopsy modality (mentioned below) that is felt best suited to obtain a tissue sample. Confirmation of the bronchoscope proximity to the SPN is substantiated via fluoroscopy and radial EBUS (rEBUS). In the Archimedes Access Kit there was an 18ga FleXNeedle, a dilation balloon and a sheath that could be utilized for bronchoscopic transparenchymal nodule access (BTPNA) which was only performed for 3 cases in our retrospective chart review. This entailed locating point of entry (POE) in the bronchus wall followed by balloon dilation where a radiopaque sheath was introduced through the lung parenchyma to access the target SPN.

### Study Procedure

Data collected included demographic information, pulmonary function testing, CT findings which was later reviewed by radiology including: extent of emphysema, extent of interstitial lung disease (ILD), bronchus sign (A-bronchus leading directly to SPN, B-bronchus visible but does not lead directly to SPN, C-there is no visible bronchus leading to SPN), generation of closest airway to the nodule, nodule features (ground glass or solid, spiculated, cavitary), nodule size, and proximity of the nodule to the pleura.^15^ Multiple thoracic radiologist assessed the severity of emphysema and ILD on CT and categorized it from mild to severe. Additionally, data collected on if patients underwent fluorodeoxyglucose-position emission tomography (FDG-PET) scans and if SPNs were avid based on standardized uptake values (SUV).

For the procedure data on biopsy technique was collected: transbronchial biopsy (TBBx), fine needle aspiration (FNA), brushing, bronchoalveolar lavage (BAL) and if linear and radial was EBUS performed. For all cases in-room fluoroscopy was used to confirm bronchoscope proximity to the SPN. Rapid on-site cytopathological examination (ROSE) was performed during the majority of cases. Cytopathology reports were reviewed for adequate tissue sample (e.g. biopsy yield) along with microbiology. Peri-procedural complications were reviewed.

### Study Outcomes

The primary endpoints were peri-and post-procedural complications such as the pneumothorax, bleeding, respiratory failure and/or admission post-procedure. Bleeding was determined to be significant if required the use of fogarty balloon inflation to tamponade bleeding, blood transfusion or admission, besides transient wedging of bronchoscope and/or instillation of cold saline or diluted epinephrine (1:10,000, 1mg/ml). For haemorrhage a grading system was applied (1-4): from suctioning of blood to requiring new admission to intensive care unit (ICU) or bronchial artery embolization; this was previously defined by Folch et al.^16^

A secondary endpoint was biopsy yield which was defined as number of patients with at least one biopsy sample sufficient for a tissue diagnosis divided by total number of patients sampled. In cases with an indeterminate (e.g. inadequate tissue sample, inflammatory cells, atypical cells, reactive bronchial cells, etc) or benign diagnosis further chart review was performed to see if any additional imaging showed an increase in SPN size or additional procedural testing (TTNA, surgical resection or repeat bronchoscopy) revealed a malignant diagnosis in order to determine diagnostic yield. An additional secondary endpoint was diagnostic yield after completing chart review for SPNs who had initially had a non-specific or benign diagnosis after an appropriate 1-year follow-up period.

### Statistical Analysis

Continuous variables were summarized as mean, standard deviation, and median (interquartile and range). Categorical variables were summarized by counts and percentages. Biopsy yield was defined as at least 1 tissue sample with adequate tissue to make a diagnosis divided by number of patients who underwent bronchoscopy. Diagnostic yield was defined as true positive (malignant at time of initial bronchoscopy and positive on repeat imaging for malignancy) divided by the total number of patients who underwent bronchoscopy for SPN(s). In order to do so any patients who were lost to follow-up and did not have any additional documentation in the EMR were exclude from final diagnostic yield analysis.

Univariate analysis with Chi-square test for categorical variables and two-sample t-test was performed to determine the impact of the collected clinical factors on diagnostic yield. Additional univariate analysis was performed for patients with non-specific diagnosis who did not have appropriate follow-up. This assumed that both the initial diagnosis was correct versus incorrect to determine the maximum and minimum sensitivity, respectively. The sensitivity was calculated based on biopsy yield definition and included patients who had inadequate follow-up. All statistical analyses were performed using SPSS software (IBM Corp., Armonk, NY). Clinical factors impact on diagnostic yield was considered statistically significant if p<0.05.

## Results

### Patient Population

Of the 96 patients who underwent VBN, 110 SPNs were biopsied; 16 patients had a diagnosis of benign/non-diagnostic and were lost to follow-up and did not have additional testing performed. This equates to a total of 18 nodules that were lost to follow-up as 1 patient had 3 nodules biopsied. Mean age was 67 years-old with equal distribution of male and female patients. A majority of patients (92%) had some form of emphysema with nearly 60% having moderate-severe emphysema based on radiographic reading along with 17% having interstitial lung disease, refer to Table 1. Pulmonary function testing was notable for a mean forced expiratory volume in 1-second (FEV1) of 61% predicted (+/-26%), mean forced vital capacity (FVC) of 84% predicted (+/-21%), FEV1%/FVC% of 71% predicted (+/-25%), mean residual volume (RV) of 129% predicted (+/-62%) and mean diffusing capacity for carbon monoxide (DL_CO_) of 48% predicted (+/-20%). The mean 6-minute walk distance was 281 meters and 29% of patients required supplemental O_2_, (refer to Table 1).

**Table 1:** Patient Demographics

|  |  | Initial Malignant Diagnosis<br>And Benign Diagnosis with Follow-up (n=80)<sup>1</sup> |  |  |
| --- | --- | --- | --- | --- |
| Variable | Total<br>(n=96) | Incorrect Diagnosis<br>(n=23) | Correct Diagnosis<sup>2</sup><br>(n=57) | p-value |
| Age, Years- Mean (SD) | 67 (10) | 65 (11) | 68 (8) | 0.11 |
| Female, N (%) | 50 (52) | 14 (61) | 27 (47) | 0.27 |
| Male, N (%) | 46 (48) | 9 (39) | 30 (53) |  |
| FEV1% - Mean (SD) | 61 (26) | 62 (27) | 60 (25) | 0.78 |
| FVC% - Mean (SD)<sup>3</sup> | 84 (21) | 84 (25) | 85 (19) | 0.94 |
| FEV1%/FVC%-Mean (SD) | 71 (25) | 70 (23) | 72 (25) |  |
| RV% - Mean (SD) | 128 (60) | 116 (59) | 135 (63) | 0.23 |
| DL<sub>CO</sub>% - Mean (SD) | 48 (21) | 50 (19) | 46 (20) | 0.38 |
| TLC%, N<sup>4</sup> | 74 | 22 | 52 | 0.49 |
| Mean (SD) | 99 (22) | 96 (19) | 100 (24) |  |
| Emphysema, N (%)<sup>5</sup> |  |  |  | 0.82 |
| None | 8 (8) | 1 (4) | 5 (9) |  |
| Mild | 34 (35) | 9 (39) | 17 (30) |  |
| Moderate | 28 (29) | 7 (30) | 19 (33) |  |
| Severe | 26 (27) | 6 (26) | 16 (28) |  |
| Interstitial Lung Disease N (%)<sup>6</sup> |  |  |  | 0.82 |
| None | 79 (83) | 19 (83) | 47 (81) |  |
| Mild | 11 (12) | 3 (13) | 6 (11) |  |
| Moderate | 3 (3) | 1 (4) | 2 (4) |  |
| Severe | 2 (2) | 0 (0) | 2 (4) |  |
| Oxygen, N (%) |  |  |  | 1.00 |
| Yes | 27 (30) | 7 (33) | 18 (33) |  |
| 6MW, M- Mean (SD) | 281 (92) | 245 (107) | 296 (76) | 0.04 |

1-Statistical analysis performed on 80 patients who had complete follow-up for non-diagnostic/benign VBN cases. 16 patients excluded for no follow-up data 1-year post VBN if benign/non-diagnostic.
2- Correct v. Incorrect Diagnosis represents the patients who had a benign/non-diagnostic at time of VBN and same diagnosis after further workup compared to patients who upon further workup was positive for malignancy (located in the incorrect column).
3-Forced vital capacity (FVC%) predicted
4- Total Lung Capacity (TLC)% is based on 80 patients who had complete follow-up
5-Emphysema severity: mild, moderate and severe based on radiographical grading as per Radiologist
6-ILD severity: mild, moderate and severe based on radiographical grading as per Radiologist

### Nodule Characteristics and Bronchoscopic Features

Nodules were visible on 62% of cases on chest radiograph and/or fluoroscopy and bronchus sign was present for 85% of cases (either direct bronchus leading to nodule or visible airway on cross section CT imaging). There were similar distributions of SPNs in both the right upper (RUL) and left upper lobes (LLL), 34% and 36%, respectively. Of SPNs biopsied, 81% were solid with the remainder either a ground glass opacity or mixture of both. The mean nodule size based off major axis (longest diameter) was 23 mm. For other nodule characteristics refer to Table 2.

**Table 2:** Nodule Characteristics

|  |  | **Initial Malignant Diagnosis**<br>**And Benign Diagnosis with Follow-up (n=80)<sup>1</sup>** |  |  |
| --- | --- | --- | --- | --- |
| **Variable** | **Total Patients (n=96)** | **Incorrect Diagnosis**<br>**(n=23)** | **Correct Diagnosis**<br>**(n=57)** | **p-value** |
| Nodule Size (mm)<sup>2</sup> |  |  |  | 0.53 |
| Mean (SD) | 23 (13) | 25 (14) | 23 (12) |  |
| <10mm | 6 (3) | 5 (3) | 6 (3) |  |
| 11-20mm | 14 (3) | 14 (3) | 15 (3) |  |
| 21-30mm | 25 (3) | 24 (2) | 25 (3) |  |
| >31mm | 39 (7) | 34 (2) | 39 (7) |  |
| Visible on CXR, N (%) |  |  |  | 0.08 |
| Yes | 57 (62) | 11 (48) | 38 (69) |  |
| Bronchus Sign, N (%)<sup>3</sup> |  |  |  | 0.59 |
| C | 13 (14) | 2 (9) | 9 (16) |  |
| B | 30 (32) | 7 (30) | 19 (34) |  |
| A | 50 (54) | 14 (61) | 28 (50) |  |
| Artery Sign, N (%) |  |  |  | 0.50 |
| Yes | 90 (97) | 22 (96) | 55 (98) |  |
| Spiculated Margin?, N (%) |  |  |  | 0.30 |
| Yes | 78 (84) | 18 (78) | 49 (88) |  |
| Calcification, N (%) |  |  |  | 0.83 |
| Yes | 7 (8) | 2 (9) | 4 (7) |  |
| Cavitation, N (%) |  |  |  | 0.77 |
| Yes | 9 (10) | 2 (9) | 6 (11) |  |
| Proximity from pleura<br>(mm), N | 75 | 19 | 44 | 0.98 |
| Mean (SD) | 15.4 (15) | 14.3 (13) | 14.4 (15) |  |
| PET Avid (0-n 1-y), N (%) |  |  |  | 0.96 |
| Yes | 83 (97) | 22 (96) | 47 (96) |  |
| SUV Uptake, N | 73 | 20 | 42 | 0.67 |
| Mean (SD) | 9 (8) | 9 (8) | 10 (9) |  |

1-Statistical analysis performed on 80 patients who had complete follow-up for non-diagnostic/benign VBN cases.
2-Nodular size based off largest diameter of the x/y dimensions.
3- C-denotes no visible bronchus leading to SPN, B-represents visible bronchus in same cross section CT image as SPN but does not go directly to SPN, A-denotes bronchus leading directly to SPN

Eight-four percent of cases had only one SPN biopsied. In most cases Linear EBUS was performed first if pet-avid nodules were noted on imaging. Tunneling was only performed in the 3 BTPNA cases. The most common sampling modality was brushing in 92% of cases; 54% had FNA and 59% had TBBx performed. Radial-EBUS was commonly used to help identify lesion (94%) with a similar distribution of eccentric or concentric views (Table 3).

**Table 3:**
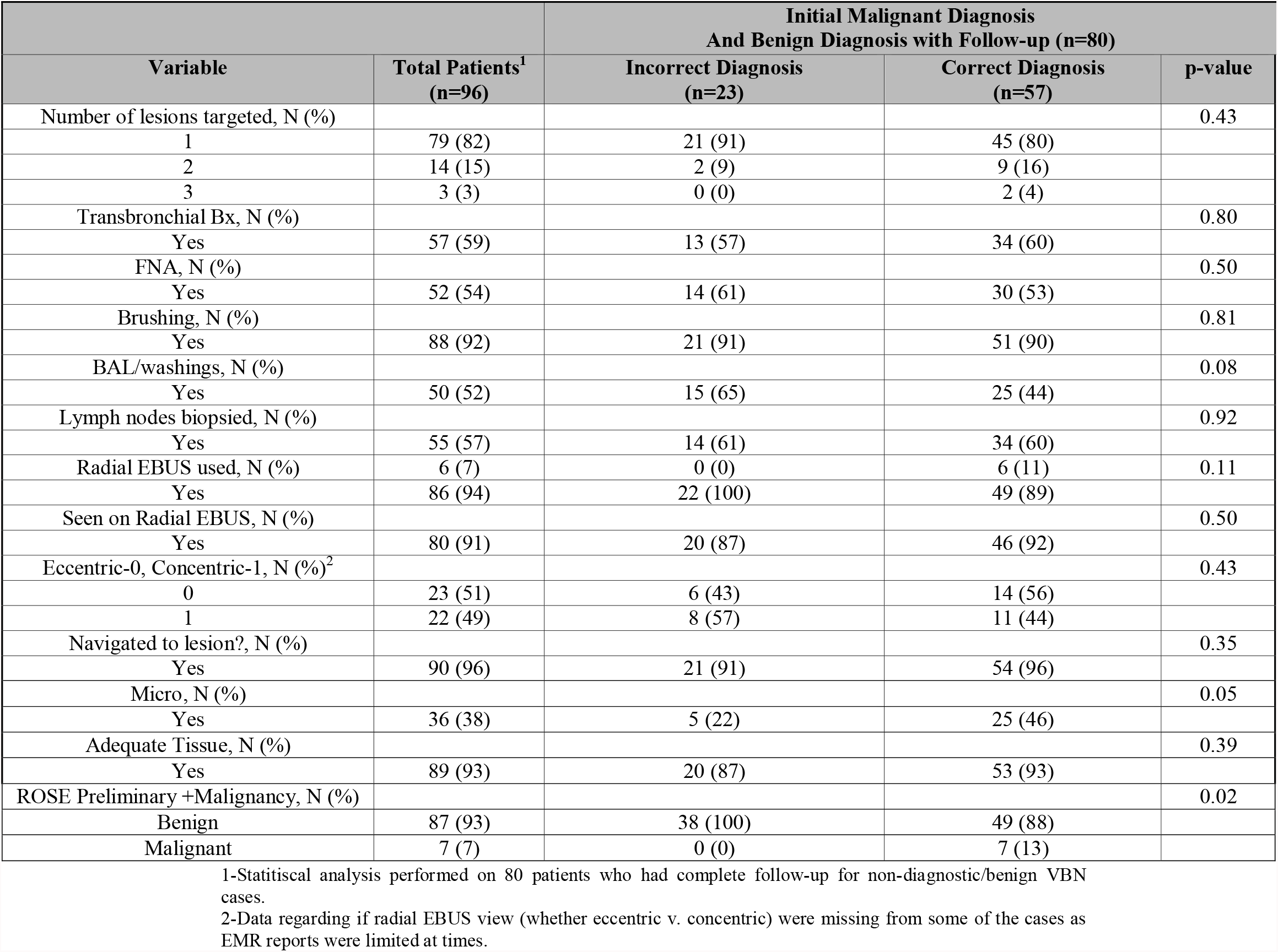
Virtual Bronchoscopic Navigational Features

Diagnoses from bronchoscopy were 38% benign, 24% malignant (23 nodules), 7% non-diagnostic and 31% of cases were infectious/benign. Follow-up CT imaging was performed for the 40 patients with benign (including infectious) and non-diagnostic SPNs within one year of initial bronchoscopy. The remaining cases (16 patients) who had a benign or were non-diagnostic were lost to follow-up. Repeat bronchoscopy was performed in 2% of non-diagnostic cases, 20% underwent surgical resection for highly suspicious nodules and 11% underwent CT-guided TTNA for further diagnostic workup, refer to Table 4

**Table 4:** Diagnostic Yield after Appropriate Work-up with Initial Benign/Non-Diagnostic

| Variable | Total Patients<sup>1</sup><br>(n=96) | Diagnosis with Follow-up |  | p-value |
| --- | --- | --- | --- | --- |
|  |  | Incorrect Dx<br>(n=39) | Correct Dx<br>(n=57) |  |
| Bronchoscopy Diagnosis, N (%) |  |  |  | <.0001 |
| Benign | 36 (38) | 26 (67) | 10 (18) |  |
| Malignant | 23 (24) | 0 (0) | 23 (40) |  |
| Non-Diagnostic<sup>2</sup> | 4 (4) | 2 (5) | 2 (4) |  |
| Infectious/Benign | 30 (31) | 10 (26) | 20 (35) |  |
| Infectious/Inadequate<sup>3</sup> | 3 (3) | 1 (3) | 2 (4) |  |
| Final diagnosis different from bronchoscopy, N (%) |  |  |  | <.0001 |
| No | 57 (71) | 0 (0) | 57 (100) |  |
| Yes | 23 (29) | 23 (100) | 0 (0) |  |
| Secondary Testing, N (%)<sup>4</sup> |  |  |  | <.0001 |
| None | 16 (21) | 15 (39) | 1 (3) |  |
| Repeat VNB | 2 (3) | 1 (3) | 1 (3) |  |
| CT-guided TTNA | 11 (15) | 7 (18) | 4 (11) |  |
| Surgical Biopsy/Resection | 15 (20) | 13 (33) | 2 (6) |  |
| Repeat CT Imaging only | 28 (37) | 3 (8) | 25 (69) |  |
| No Further Work-up Warranted | 3 (4) | 0 (0) | 3 (8) |  |
| Malignant- 2nd test, N (%) |  |  |  | <.0001 |
| No | 33 (58) | 1 (4) | 32 (97) |  |
| Yes | 24 (42) | 23 (96) | 1 (3) |  |
| Follow up CT, N (%)<sup>5</sup> |  |  |  |  |
| No | 18 (31) | 2 (18) | 4 (12) |  |
| Yes | 40 (69) | 9 (82) | 30 (88) |  |
| Repeat CT results, N (%)<sup>6</sup> |  |  |  | <.0001 |
| 0 | 31 (80) | 3 (33) | 28 (93) |  |
| 1 | 8 (21) | 6 (67) | 2 (7) |  |
| Time from bronchoscopy to final diagnosis, (Days) | 21 | 17 | 4 | 0.002 |
| Mean (SD) | 131.5 (170) | 162.5 (176) | 0 (0) |  |
| Malignant From Bronch, N (%) |  |  |  | <.0001 |
| No | 73 (76) | 39 (100) | 34 (60) |  |
| Yes | 23 (24) | 0 (0) | 23 (40) |  |
| Benign from Bronch, N (%) |  |  |  | <.0001 |
| No | 23 (24) | 0 (0) | 23 (40) |  |
| Yes | 73 (76) | 39 (100) | 34 (60) |  |
| Confirmed Benign, N (%)<sup>7</sup> |  |  |  | <.0001 |
| 0 | 23 (32) | 23 (59) | 0 (0) |  |
| 1 | 34 (47) | 0 (0) | 34 (100) |  |
| 2 | 16 (21) | 16 (41) | 0 (0) |  |
| False Benign (Later malignant), N (%) |  |  |  | <.0001 |
| No | 34 (60) | 0 (0) | 34 (100) |  |
| Yes | 23 (40) | 23 (100) | 0 (0) |  |

1-Statistical analysis performed on 80 patients who had complete follow-up for non-diagnostic/benign VBN cases.
2-Bronchoscopy diagnosis that were deemed initially non-diagnostic ended up having a correct (i.e. benign diagnosis) based on further workup that the SPN was benign.
3-Bronchoscopy diagnosis was deemed infectious/inadequate if the sample quality was poor and +microbiology was not considered an adequate explanation for the SPN present.
4-For secondary testing it was performed if diagnosis of malignancy was not confirmed on the initial bronchoscope, in some cases none was performed due to the patient being lost to follow-up or being seen at another center where we did not have access to their EMR records.
5-Patients who were deemed to have an incorrect diagnosis based off on initial bronchoscopy not always went for CT but rather went for alternative secondary testing
6- 0-denotes no change in lesion size/resolved, 1-represents lesion has increased
7- 0-represents no and 1 represents yes, 2-denotes no further follow-up.

Overall there were 7 (7%) adverse events based on total number of patients who had some form of complication with 1-pneumothorax (PTX), 3-significant bleeding (requiring Fogarty balloon inflation or temporary hemodynamic instability with hypotension requiring procedure to be aborted) and 3-admitted for respiratory failure, refer to Table 5. Only 1 of the patients that had significant bleeding required admission post-procedure for monitoring. Based on intervention required for bleeding the patients met grade 3 as per Folch et al. bleeding severity score.^16^

**Table 5:**
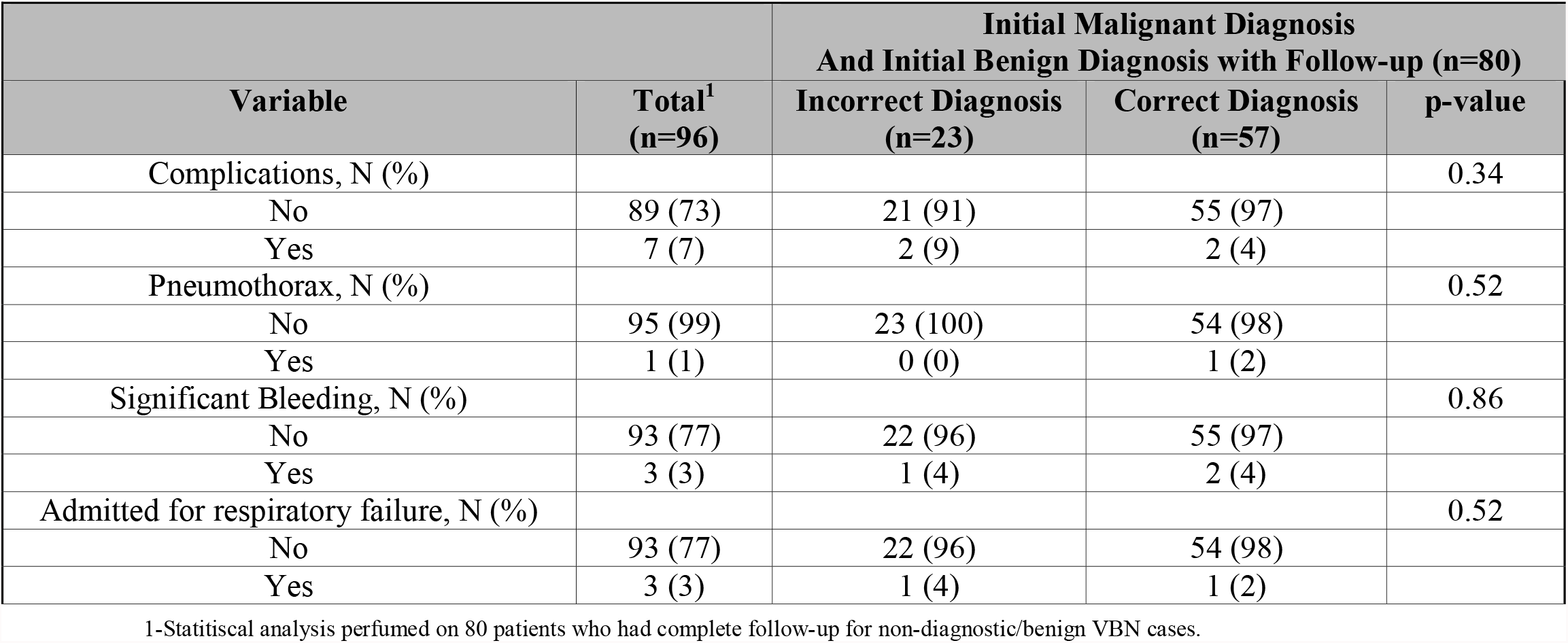
Peri-Operative Complications

|  |  | **Initial Malignant Diagnosis**<br>**And Initial Benign Diagnosis with Follow-up (n=80)** |  |  |
| --- | --- | --- | --- | --- |
| **Variable** | **Total<sup>1</sup>**<br>**(n=96)** | **Incorrect Diagnosis**<br>**(n=23)** | **Correct Diagnosis**<br>**(n=57)** | **p-value** |
| Complications, N (%) |  |  |  | 0.34 |
| No | 89 (73) | 21 (91) | 55 (97) |  |
| Yes | 7 (7) | 2 (9) | 2 (4) |  |
| Pneumothorax, N (%) |  |  |  | 0.52 |
| No | 95 (99) | 23 (100) | 54 (98) |  |
| Yes | 1 (1) | 0 (0) | 1 (2) |  |
| Significant Bleeding, N (%) |  |  |  | 0.86 |
| No | 93 (77) | 22 (96) | 55 (97) |  |
| Yes | 3 (3) | 1 (4) | 2 (4) |  |
| Admitted for respiratory failure, N (%) |  |  |  | 0.52 |
| No | 93 (77) | 22 (96) | 54 (98) |  |
| Yes | 3 (3) | 1 (4) | 1 (2) |  |

1-Statistical analysis performed on 80 patients who had complete follow-up for non-diagnostic/benign VBN cases.

### Biopsy Yield and Final Diagnostic Yield

Based on adequate tissue sample (from cytopathology report) the overall biopsy yield was 93%, Table 3. Over one-year of follow-up other testing for non-diagnostic or benign/infectious SPNs revealed an additional 23 (42%) patients diagnosed with malignancy and were therefore “false-negative” at time of bronchoscopy, with the remainder of the 33 initially benign/non-diagnostic biopsies (58%) remaining negative, refer to Table 4. There were 16 patients that were excluded from statistical analysis for the final one-year diagnostic yield based on the definition from AQuIRE and NAVIGATE trials due to being lost to follow-up. There was a final diagnostic yield of 72%, refer to Table 4.^12,13^ There was a statistically significant difference in diagnostic yield in regards to the 6-minute walk distance with a lower walk-distance in those with false-negative diagnoses (245 meters +/-88 meters) than in those with correct diagnoses (296 meters +/-76 meters) (p=0.039).

### Minimum and Maximum Sensitivity

In the AQuIRE trial for patients who did not have complete follow-up for benign or non-diagnostic SPN on original VBN they were able to determine sensitivity based on the assumption of all the indeterminate SPNs that were lost to follow-up as all “false-negative” (minimum sensitivity) or all “true-negative” (maximum sensitivity).^12^ Minimum sensitivity of all 96 patients who underwent VNB was 59.3% (p<0.0001) with a maximum sensitivity of 76% (p<0.0001), refer to Supplemental Table 1.

## Discussion

Our study was a retrospective review of 96 patients that underwent VBN utilizing the Archimedes platform from December 2016 to October 2019. In this study the goal was to evaluate the biopsy and diagnostic yield and safety profile for VBN in patients with underlying advanced lung disease. In our patient population 92% had some form of emphysema and 15% had ILD; overall 30% were on supplemental oxygen.

The overall complication rate was 7% with 1 patient having PTX (1%), 3 patients having significant bleeding (3%) and 3 patients admitted for respiratory failure (3%) during the peri-procedure period. Comparing to Ost et al. (AQuIRE trial) there were 10 patients who developed pneumothorax (2% of total sample size) after undergoing VBN.^12^ Our study and AQuIRE trial are comparable to a recent systemic review and meta-analysis by Han et al. that included 426 patients that demonstrated an overall rate of PTX 2% with less than 1% requiring chest tube placement and less than 1% having hemoptysis.^17^

When comparing VBN versus TTNA the risk of developing PTX is higher in the TTNA group. A recent meta-analysis showed the rate of PTX ranged from 7%-50% in TTNA with approximately 6% requiring some form of intervention which was not specified.^18^ In patients with advanced lung disease (e.g., COPD, ILD, combined pulmonary fibrosis and emphysema (CPFE), etc.) Yoon et al. demonstrated there is an increase odds ratio (OR) of 1.58 (p<0.001) to develop pneumothorax with fluoroscopic or CT-guided TTNA with a tendency to require chest tube insertion with a OR of 2.75 (p<0.001) which was again confirmed by Kolu et al with an OR of 4.62 (p<0.001) for emphysematous patients undergoing TTNA to develop pneumothorax.^19,20^ These 2 studies demonstrate the need for an acceptable alternative means for sampling SPNs besides TTNA in patients with advanced underlying lung disease with risk of pneumothorax. In our study we demonstrated VBN is a safe modality compared to TTNA when sampling SPNs in emphysematous patients.

The question lies does VBN provide a similar diagnostic yield to TTNA, the gold standard when biopsying SPNs? There was recently a large retrospective chart review in Korea by Lee et al. of 9384 TTNA that showed an overall diagnostic yield of 91% with a sensitivity and specificity of 93% and 87%, respectively.^**21**^ Compared to our diagnostic yield of 72% with a maximum sensitivity of 76% which is on par with NAVIGATE trial of 73%.^13^ Despite a consistent diagnostic yield in the trials mentioned a previous metanalysis of 39 studies show much heterogeneity with a yield range of 46% to 86%; possibly contributing to prospective studies and VBN performed outside of research setting often having a lower yield.^**12**,**22**,**23**^ Currently, the yield for VBN despite different platforms being utilized are maxed 70-80% range when biopsying SPNs.

### Benefits/Limitations

Nearly all VBNs that was performed at TUH was accomplished by a single operator alongside anesthesiologists who were accustomed to treating to patients with advance lung disease therefore limiting generalizability to routine clinical practice which may potentially overestimated the final diagnostic yield and limit adverse events that would normally be seen by this population. Additional limitation was our study was retrospective chart review dependent on EMR documentation which at times was missing information; specifically, incomplete three-dimensional measurements of SPNs, difficulty in discerning what form of biopsy technique was used for each case, if there was adequate tissue sample based off the cytopathology reports and follow-up post procedure especially if the VBN was diagnostic was limited (e.g. CT imaging and secondary testing). Additionally, we did not factor in how much of the diagnostic yield can be attributed to rEBUS versus VBN by itself. Also, with TUH not being a cancer center but instead having a large referral base, patients commonly seek care at alternative centers which resulted in incomplete follow-up for 16 patients which potentially impacted final diagnostic yield. Potential benefit was the application of our retrospective study to the general public consider TUH is a university hospital without a connecting cancer institute the percentage of lung cancer diagnosis would be theoretically higher compared to what was seen at our institution.

Our study albeit retrospective, single center had the ability to demonstrate VBN is a safe modality to perform on patients with advance lung disease when biopsy SPNs with minimal risk of PTX compared to TTNA where the risk can range anywhere from 7-50%.^18^ The limitation of VBN compared to TTNA even with rEBUS is the diagnostic yield has been repeatedly shown to be in the 70-80% range versus TTNA with a yield range of 77-98%.^17^ With future development of robotic bronchoscopic platforms it will be interesting to see if the diagnostic yield will improve with biopsying SPNs and possibly be an alternative to TTNA.

## Supporting information

Table S1-Min/Max Sensitivity

## Data Availability

All data produced in the present work are contained in the manuscript

## Abbreviations

(SPNs): Suspicious Pulmonary Nodules
(VBN): Virtual Bronchoscopic Navigation
(ILD): Interstitial Lung Disease
(TUH): Temple University Hospital
(PFT): Pulmonary Function Testing
(FEV1): Forced Expiratory volume in 1-second
(FVC): Force Vital Capacity
(DL_CO_): Diffusing Capacity for Carbon Monoxide
(TLC): Total Lung Capacity
(LDCT): Low Dose Computed Tomography
(NLST): National Lung Screening Trial
(VATS): Video-Assisted Thoracic Surgery
(TTNA): Transthoracic Needle Aspiration
(EBUS): Endobronchial Ultrasound
(ENB): Electromagnetic Navigational Bronchoscopy
(rEBUS): radial EBUS
(BTPNA): Bronchoscopic Transparenchymal Nodule Access
(PTX): Pneumothorax
(FDG-PET): Fluorodeoxyglucose-Position Emission Tomography
(TBBx): Transbronchial Biopsy
(FNA): Fine Needle Aspiration
(BAL): Bronchoalveolar Lavage
(ROSE): Rapid On-site Cytopathological Examination
(ICU): Intensive Care Unit
(RUL): Right Upper Lobe
(LUL): Left Upper Lobe
(CPFE): Combine Pulmonary Fibrosis and Emphysema
(OR): Odds Ratio

