## Supplementary material for "Safety and Efficacy of Navigational Bronchoscopy with Archimedes Virtual Bronchoscopic Platform in Patients with Advanced Pulmonary Disease": Table S1-Min/Max Sensitivity: Table S1-Min:Max Sensitivity.docx

Table S1: Minimum and Maximum Sensitivity

1-Assuming all 16 patients lost to follow-up had the wrong diagnosis.

2-Assuming all 16 patients lost to follow-up had the correct diagnosis.

|  | | **Diagnosis with Follow-up** | | |  |
| --- | --- | --- | --- | --- | --- |
| **Variable** | **Total (n=96)** | | **Incorrect Diagnosis (n=39)** | **Correct Diagnosis (n=57)** | **p-value** |
| Final diagnosis different from bronchoscopy, N (%)^1^ |  | |  |  | <.0001 |
| No | 73 (76) | | 16 (41) | 57 (100) |  |
| Yes | 23 (24) | | 23 (59) | 0 (0) |  |
| Final diagnosis different from bronchoscopy, N (%)^2^ |  | |  |  | <.0001 |
| No | 73 (76) | | 0 (0) | 73 (100) |  |
| Yes | 23 (24) | | 23 (100) | 0 (0) |  |
